# Regular snoring is associated with uncontrolled hypertension: A longitudinal objective assessment of nightly snoring and blood pressure

**DOI:** 10.1101/2023.03.02.23286654

**Authors:** Bastien Lechat, Ganesh Naik, Sarah Appleton, Jack Manners, Hannah Scott, Duc Phuc Nguyen, Pierre Escourrou, Robert Adams, Peter Catcheside, Danny J Eckert

## Abstract

**Background:** Snoring may be a risk factor for cardiovascular disease and stroke. However, most prior studies have relied on subjective snoring evaluation from self-reports, or relatively short time-scale objective measures in small samples. This study assessed the prevalence of objectively quantified snoring measured over multiple months, and its association with blood pressure and hypertension.

**Methods:** 12,287 participants were monitored nightly in-home for approximately six months using an under-the-mattress sleep sensor to estimate the average percentage of sleep time spent snoring per night and the apnea-hypopnea index (AHI). Blood pressure cuff measurements from multiple daytime assessments were averaged to define uncontrolled hypertension based on mean systolic blood pressure ≥140 mmHg and/or a mean diastolic blood pressure ≥90 mmHg. Associations between snoring and uncontrolled hypertension were examined using logistic regressions controlled for age, body mass index (BMI), sex, and AHI.

**Findings:** Participants were predominantly middle-aged (mean±SD; 50±12 y) and male (88%). There were 2,467 cases (20%) with uncontrolled hypertension. Approximately 29, 14 and 7% of the study population snored for an average of >10, 20, and 30% per night, respectively. A higher proportion of time spent snoring (75th vs. 5th; 12% vs. 0·04%) was associated with an ∼1·9-fold increase (OR [95%CI]; 1·87 [1·63, 2·15]) in uncontrolled hypertension independent of sleep apnea. The effect size of the association between snoring and uncontrolled hypertension was higher in younger adults and those who were not obese (BMI <30 kg/m^2^).

**Interpretations:** Multi-night recordings in a large consumer sample indicate that snoring is common, and that snoring duration is positively associated with hypertension. These findings highlight the potential clinical utility of simple, objective, and non-invasive methods to detect snoring.

**Funding Source:** This was an unfunded, investigator-initiated study led by the Adelaide Institute for Sleep Health sleep research team. DJE is supported by a National Health and Medical Research Council (NHMRC) of Australia Leadership Fellowship (1196261).

**Research in context:** *Evidence before this study:* We searched PubMed, Google, and Google Scholar for research articles published in English up to June 1, 2022, using common search terms including “wearable”, “nearable”, “sleep apnea”, and “snoring”. Articles were also retrieved through searching the citations of known literature. Snoring is a major feature of sleep disordered breathing, including hypopnea events which reflect partial airway obstruction typically with snoring. A meta-regression of 63 studies reported a highly variable snoring prevalence of between 2-83% in men and between 1-71% in women. These studies relied on self-reported snoring assessments, which may not be sufficiently reliable to evaluate prevalence and potential snoring impacts on cardiovascular health outcomes. A few small scale clinical and epidemiological studies with objective snoring assessments on a single night suggest associations between snoring and adverse cardiovascular health outcomes. However, a recent meta-analysis indicates that assessment of sleep apnea severity is highly variable night to night. Like sleep apnea severity, single time-point estimation of snoring parameters may not reliably reflect potentially problematic features of snoring and cumulative exposure risks over time.

*Added value of this study:* We investigated the prevalence of snoring and its association with uncontrolled hypertension on the largest dataset to date (>12,000 participants), including multi-night assessment of snoring over ∼6 months (∼2 million nights in total). Approximately 15% of the study population snored for an average of 20% per night, and a higher proportion of time spent snoring was associated with an 87% increase in uncontrolled hypertension independent of sleep apnea severity.

*Implications of all the available evidence:* These findings provide important insight into the consequences of snoring on hypertension risk and highlight the potential need to consider snoring as part of clinical care and management of sleep problems. These findings demonstrate the potential clinical utility of simple, objective, and non-invasive methods to detect and evaluate snoring.

## Introduction

Snoring is common and reflects narrowing and tissue vibration of upper airway soft tissues and structures ^1^. The prevalence of habitual loud snoring in the general population remains unclear and is challenging to define in the absence of agreed standards for snoring assessments, knowledge regarding clinical impacts of snoring itself, and a reliance, in most existing epidemiological studies, on self- or bed partner-reports of snoring frequency and/or intensity. For example, a meta-regression of 63 studies reported a snoring prevalence between 2-83% in men and between 1-71% in women^2^. Snoring prevalence in clinical populations is also unclear. Snoring is a major feature of sleep disordered breathing, including hypopnea events which reflect partial airway obstruction typically with snoring. Consequently most, but not all, individuals with obstructive sleep apnoea (OSA) frequently snore loudly on most nights. However, based on self-report snoring assessments in one of the largest epidemiological studies of sleep disordered breathing effects on health, the Sleep Heart Health Study (SHHS), one third of participants with OSA did not report any snoring while one third of participants who were snorers did not meet criteria for OSA^3^. However, self-reported snoring assessments may not be sufficiently reliable to evaluate potential snoring impacts on cardiovascular health outcomes.

Physiological consequences of partial upper airway obstruction and snoring include substantial negative intrathoracic pressure swings that increase transmural pressure pre- and after-loads on the heart, and blood pressure surges associated with brief awakenings from sleep (cortical arousals). Tissue vibrations themselves may also be injurious to upper airway and surrounding tissues, including the carotid arteries^4,5^.

Acknowledging the shortcomings of prior attempts to quantify snoring and the need for more systematic and objective assessments of snoring to evaluate potential ill-effects of snoring on cardiovascular health outcomes, snoring has been associated with multiple sub-clinical markers of cardiovascular pathology. This includes elevated blood pressure^6-8^, increased carotid-intima-media thickness^9^, stenosis^10^, and atherosclerosis^11^. These effects could partly reflect mechanical stress imposed by snoring vibrations on the upper airway in combination with a range of shared risk factors for OSA and cardiovascular disease, such as obesity and a sedentary lifestyle. In addition, snoring sound pressure levels are often substantially higher than indoor night time noise levels recommended by the World Health organization^12^, and comparable to levels associated with poor cardiovascular health, including high blood pressure^12,13^. Thus, loud snoring may interfere with recuperative sleep and contribute to hypertension risk and other adverse outcomes in snorers and their bedpartners through noise disturbance effects. Snoring could therefore be an important risk factor for hypertension and incident cardiovascular events, as recently shown with incident stroke in the SAVE trial^14^. However, in other studies snoring was not found to be associated with all-cause mortality, or incident cardiovascular event or stroke^15,16^. Thus, the current evidence is conflicting.

Furthermore, large night-to-night variability in markers of OSA severity, such as the apnea-hypopnea-index, is now well established^17-19^. While no studies have examined night-to-night variability in objective snoring assessments, given mechanistic overlap between hypopneic events and snoring, and night-to-night variability in sleep depth and posture, single night snoring assessments may also not reliably reflect typical snoring and cumulative exposure risks over time.

Therefore, existing evidence for associations between snoring and cardiovascular health outcomes remain limited and are primarily based on self-reported snoring assessments, or clinical/epidemiological samples with single night objective snoring assessments in relatively small sample sizes. This study therefore aimed to determine the prevalence of snoring and its association with hypertension using multi-night objective assessment of snoring and multiple daytime blood pressure assessments in a large population sample.

## Methods

### Participants

This study analyzed data from 12,287 participants who registered to use both an under-mattress sleep sensor (Withings Sleep Analyzer: WSA) and an FDA-registered home-blood pressure monitor between July 2020 and April 2021^20^. Further inclusion criteria were ≥28 nights of sleep and snoring recordings and at least five blood pressure measurements over the recording period for assessment of uncontrolled hypertension outcomes. The current study was approved by the Flinders University Human Research Ethics Committee (Project number: 4291).

### Snoring and blood pressure assessments

The WSA is a non-wearable sleep monitoring device placed under-the-mattress that detects snoring and estimates the apnea-hypopnea index (AHI) and sleep stages. This is achieved via automated proprietary algorithms from a built in microphone and ballistograhic assessment of movement, heart rate and respiratory motion from a pressure sensor^21^. Features characteristic of the power spectrum of snoring sound are calculated and snoring is scored over 1-minute epochs. To be classified as a snoring epoch, ≥ 50% of the epoch must contain snoring. A post-processing window over 3 consecutive 1 min epochs is then applied to define snoring. This has the advantage of limiting the number of false positives due to extraneous noise. Snoring duration is then expressed as a percentage of total sleep time by dividing snoring time by the estimated total sleep time (hereafter termed “snoring duration”).

Blood pressure values were obtained via Withings blood pressure monitor measurements undertaken by each participant. The home-blood pressure monitor comes with an instruction booklet that outlines how to take a blood pressure measurement, and the user is instructed to 1) rest for ≥5 minutes before taking a measurement, 2) be seated in a comfortable position and in a quiet area with legs uncrossed, feet flat on the floor and back/arm supported, 3) not speak during the measurements and 4) perform the measurement on their left arm. Uncontrolled hypertension was defined as a mean systolic blood pressure ≥140 mmHg or mean diastolic blood pressure ≥90 mmHg^22^ averaged across all available blood pressure measurements over the monitoring period.

### Statistical analysis

The primary exposure variables were mean AHI and mean snoring duration across all available nights of data. OSA severity categories were defined using standard clinical cut-offs^23^ (<5 events/h sleep= no OSA, ≥5 and <15= mild, ≥15 and <30= moderate and ≥30 events/h sleep= severe OSA). Snoring was categorized using quartiles and tertiles where necessary for interpretation but was otherwise kept as a continuous variable in the analyses.

The prevalence of snoring and mean snoring duration was examined separately by sex. Linear regression models controlling for age, sex, BMI and mean AHI were used to investigate the potential association between snoring duration with systolic and diastolic blood pressure. Odds ratio (ORs) and 95% confidence interval (CIs) were determined using logistic-regression models to assess the association between mean snoring duration and uncontrolled hypertension adjusted for age, sex, BMI, and average AHI.

AHI and mean snoring duration were modelled using restricted cubic splines to account for potential non-linear associations. Interactions between snoring duration with age, sex and BMI were also investigated. When interactions where significant, BMI was categorized as normal (< 25 kg/m^2^), overweight (BMI between 25 and 30 kg/m^2^) and obese (BMI ≥ 30 kg/m^2^). When interactions with age were observed, age was categorized using a median split. As a secondary analysis, snoring duration was categorized using tertiles and the contribution of snoring in each OSA category (from mean AHI) was examined using logistic regression adjusted for available confounders. Logistic and linear regression were performed in the R programming language.

#### Funding Source

This was an unfunded, investigator-initiated study led by the Adelaide Institute for Sleep Health sleep research team.

## Results

### Participant characteristics

The characteristics of the 12,287 participants included in this study are summarized in Table 1. Participants were middle aged (∼50 years), generally overweight (BMI ∼28 kg/m^2^) and predominantly male (88%). Each participant had a median [IQR] of 29 [12, 81] repeat blood pressure recordings, and mean±SD 181±69 nightly recordings of sleep and snoring throughout the study period. Approximately 45%, 29%, 14% and 7% of the study population snored for more than 5%, 10%, 20%, and 30% of the night, respectively. For men, these rates were 46%, 30%, 15%, and 7.6%, whereas for women the equivalent rates were 36%, 22%, 9%, and 4%.

**Table 1.**
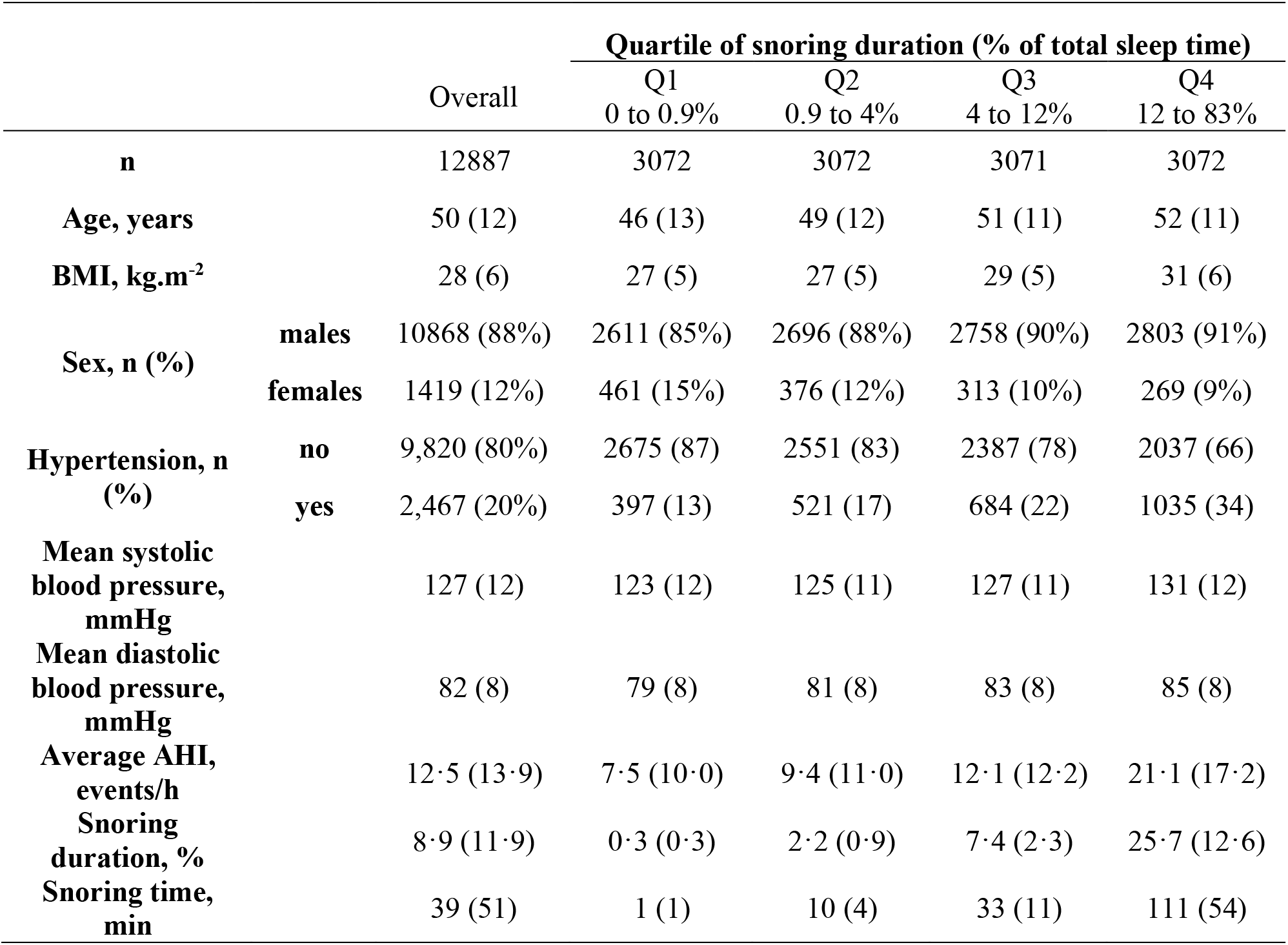
Baseline characteristics of the sample population in relation to snoring duration

The Spearman correlation coefficient between snoring duration and mean AHI was 0·42 (*p* <0·001). Participants with moderate and severe OSA exhibited substantially higher proportions of sleep time with snoring (Table 2) compared to participants with mild OSA, who were quite evenly represented in each snoring quartile. Participants without OSA had the lowest proportion of sleep time with snoring.

**Table 2.** OSA severity [n (%)] in relation to snoring duration quartiles.

|  |  | Quartiles of percent of sleep time spent snoring |  |  |  |
| --- | --- | --- | --- | --- | --- |
|  |  | 0 to 0·9% | 0·9 to 4% | 4 to 12% | 12 to 83% |
| **Obstructive**<br>**sleep apnea**<br>**severity**<br>**category<sup>1</sup>** | **No OSA** | 1727<br>(38%) | 1413<br>(31%) | 974<br>(22%) | 405<br>(9%) |
|  | **Mild** | 920<br>(22%) | 1070<br>(25%) | 1250<br>(30%) | 996<br>(24%) |
|  | **Moderate** | 303<br>(13%) | 420<br>(19%) | 596<br>(26%) | 943<br>(42%) |
|  | **Severe** | 115<br>(9%) | 171<br>(13%) | 254<br>(20%) | 730<br>(57%) |

<sup>1</sup>OSA category was defined using standard clinical cut-offs (<5 events/h sleep= no OSA, ≥5 and <15= mild, ≥15 and <30= moderate and ≥30 events/h sleep= severe OSA)

### Snoring and blood pressure

Increased snoring duration was associated with both increased systolic and diastolic blood pressure even after adjusting for averaged AHI, age, BMI and sex (Figure 1). The association between snoring duration and blood pressure modelled without interactions is depicted in Figure

**Figure 1.**
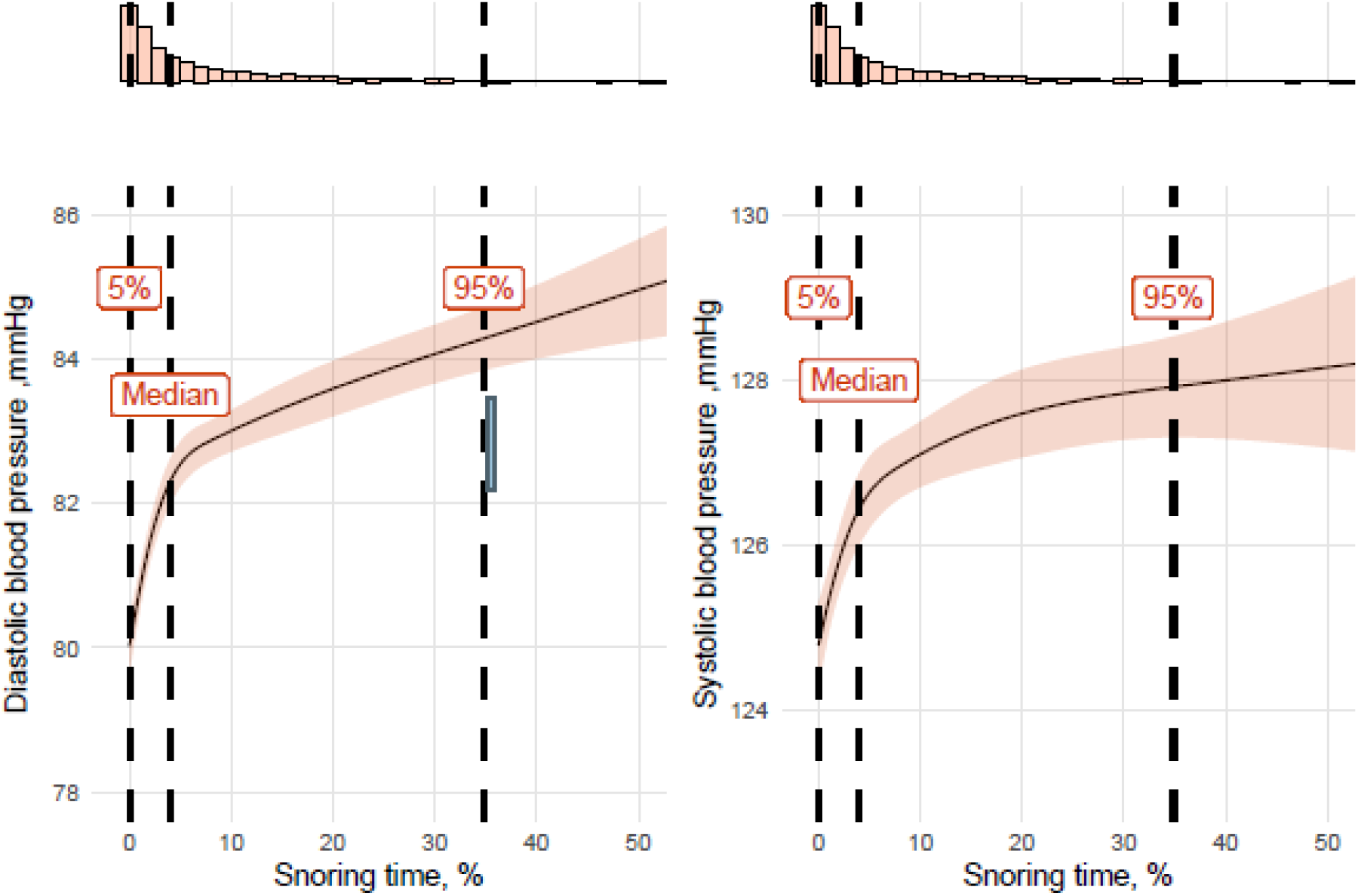
Associations between snoring duration with systolic and diastolic blood pressure, modelled using 4 knots restricted cubic spline for snoring duration.

1. These data show ∼3 and ∼4 mmHg increases in systolic and diastolic blood pressure for frequent regular snoring versus infrequent snoring independent of age, BMI, sex and mean AHI, respectively. While there were significant interactions between obese and non-obese, ≤ 50 years old vs. > 50 years old (median age) in the association between snoring duration and systolic and diastolic blood pressure, the effect size of the interaction was relatively small, with only a ∼1 to 2 mmHg difference between BMI and age categories (Appendix Figure 1 and Appendix Table 1).

Similar results were observed when OSA severity was categorized and snoring duration was divided into quartiles (Table 3). In this model, severe OSA with no snoring was associated with 3.6 mmHg and 3.5 mmHg higher systolic and diastolic blood pressure compared to no snoring or OSA. Furthermore, participants with no OSA but high snoring (quartile 4) had a 3.8 mmHg and 4.5 mmHg higher systolic and diastolic blood pressure compared to participants with no sleep apnea or no snoring. Hence, the association between severe OSA-alone and blood pressure had a similar effect size to the association between snoring-alone and blood pressure.

**Table 3.** Associations between systolic and diastolic blood pressure with OSA severity categories and quartile of snoring duration. Displayed values are β (95% CI) of the association between an OSA/snoring group with the reference group (no-OSA and lowest snoring quartile). Models were adjusted for sex, age and BMI. OSA severity categories were defined using standard clinical cut-offs of the mean apnoea-hypopnoea index (<5 = no OSA, ≥5 and < 15 = mild, ≥15 and < 30 = moderate, and ≥30 events/h sleep= severe OSA).

|  |  | Quartiles of snoring duration |  |  |  |
| --- | --- | --- | --- | --- | --- |
|  |  | 0 to 0.9% | 0.9 to 4% | 4 to 12% | 12 to 83% |
| **Systolic blood pressure** |  |  |  |  |  |
| **Obstructive sleep apnea severity category<sup>1</sup>** | **No OSA** | 0 (ref) | 1.2<br>(0.4, 1.9) | 2.4<br>(1.5, 3.2) | 3.8<br>(2.6, 4.9) |
|  | **Mild** | 1.1<br>(0.3, 2.0) | 2.1<br>(1.3, 3.0) | 2.9<br>(2.1, 3.7) | 3.6<br>(2.7, 4.4) |
|  | **Moderate** | 2.6<br>(1.3, 3.9) | 2.5<br>(1.3, 3.6) | 3.0<br>(1.9, 4.0) | 4.3<br>(3.4, 5.2) |
|  | **Severe** | 3.6<br>(1.6, 5.7) | 4.2<br>(2.5, 5.9) | 3.9<br>(2.5, 5.4) | 5.5<br>(4.5, 6.5) |
| **Diastolic blood pressure** |  |  |  |  |  |
| **Obstructive sleep apnea severity category<sup>1</sup>** | **No OSA** | 0 (ref) | 1.9<br>(1.4, 2.5) | 2.9<br>(2.3, 3.5) | 4.5<br>(3.6, 5.3) |
|  | **Mild** | 1.6<br>(1.0, 2.2) | 3.1<br>(2.5, 3.7) | 4.3<br>(3.7, 4.9) | 5.0<br>(4.4, 5.6) |
|  | **Moderate** | 3.2<br>(2.3, 4.2) | 3.0<br>(2.2, 3.9) | 4.0<br>(3.3, 4.8) | 5.4<br>(4.8, 6.1) |
|  | **Severe** | 3.5<br>(2.1, 5.0) | 3.1<br>(1.9, 4.3) | 4.5<br>(3.4, 5.5) | 6.3<br>(5.6, 7.0) |

### Snoring and uncontrolled hypertension

Snoring duration was significantly associated with uncontrolled hypertension. The interaction between snoring duration and sex was not significant (*p*= 0·34). However, there were significant interactions between snoring and age (*p*= 0·003) and snoring and BMI (*p* < 0·001), with stronger associations in participants ≤ 50 years (Figure 2) compared to those > 50 years and in participants with a BMI <30 compared to ≥ 30 kg/m^2^. In all instances, snoring was significantly associated with uncontrolled hypertension, from a 20% increased likelihood of uncontrolled hypertension in those aged >50 years and obese to a 98% increase in those aged ≤ 50 years with body weight in the normal BMI range.

**Figure 2.**
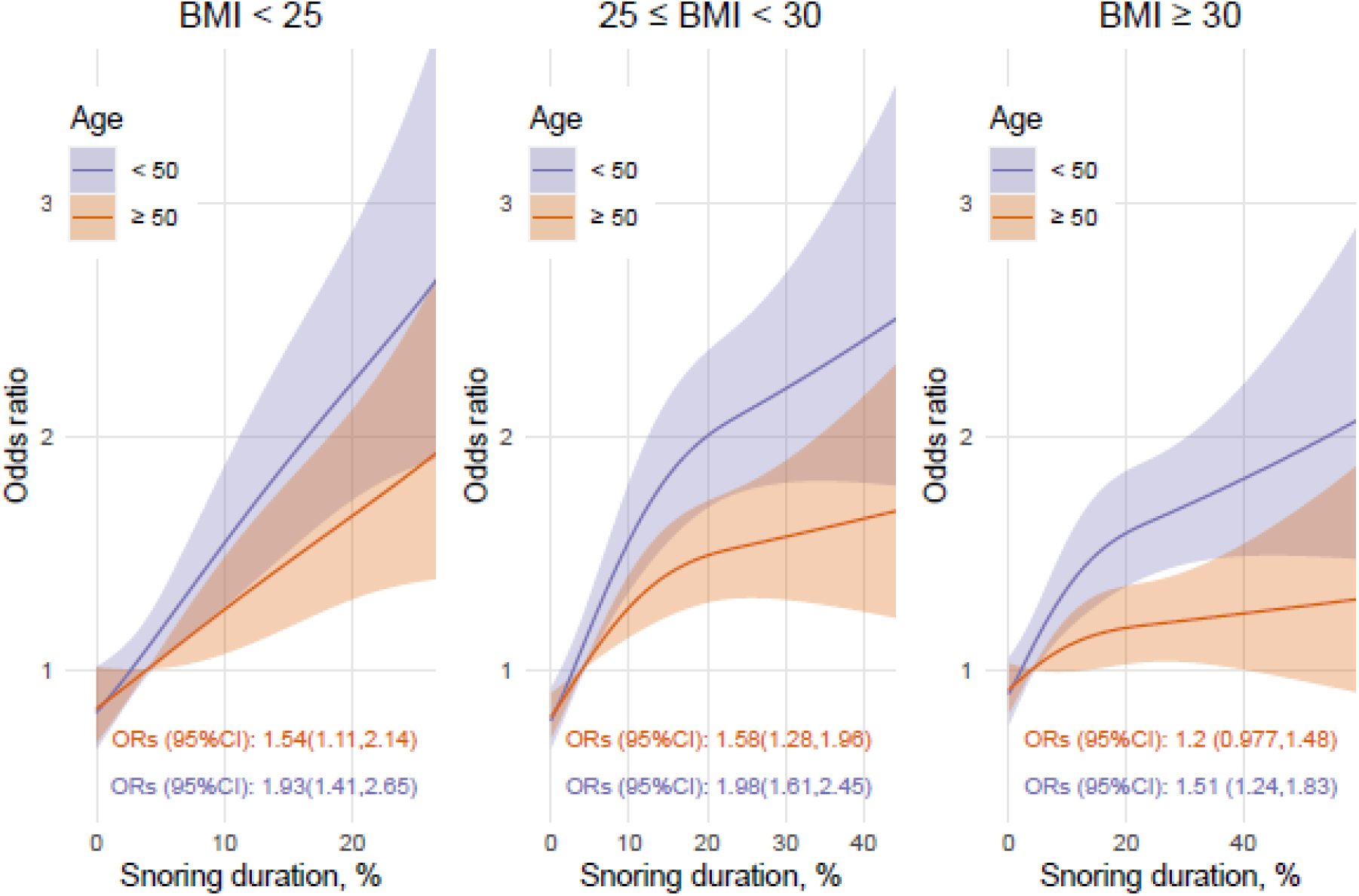
Associations between snoring duration with hypertension, modelled using 4 knots restricted cubic spline and an interaction with age categories (median split in years) and BMI categories (kg/m^2^). ORs (95%CI) represents the difference between the 5% and the 75% percent of the snoring duration distribution. Note that the 5% and 75% percent were determined separately for each BMI categories; hence slightly different x-axis.

Without considering the interactions, snoring duration was associated with an 87% increase in uncontrolled hypertension likelihood (75^th^ vs. 5^th^ percentile; 12% vs. 0·04%; OR [95%CI]; 1·87 [1.63, 2.15]). The association was non-linear and is shown in Appendix Figure 2, with a reference point using the median snoring duration. Similar results were observed when OSA severity was categorized, and snoring duration was divided into quartiles (Table 4). The association between severe OSA-alone and uncontrolled hypertension showed a similar effect size to the association between highest snoring duration-alone and uncontrolled hypertension (2.56-fold versus 2.73-fold increase).

**Table 4.** Associations between hypertension likelihood with obstructive sleep apnea (OSA) severity categories and quartile of % of sleep time spent snoring. Displayed values are ORs (95% CI) of the association between an OSA/snoring group with the reference group (no-OSA and lowest snoring quartile). Models were adjusted for sex, age and BMI.

|  |  | Quartiles of percent of sleep time spent snoring |  |  |  |
| --- | --- | --- | --- | --- | --- |
|  |  | 0 to 0.9% | 0.9 to 4% | 4 to 12% | 12 to 83% |
| Obstructive<br>sleep apnea<br>severity<br>category<sup>1</sup> | No OSA | 1 (ref) | 1.47<br>(1.17, 1.86) | 1.81<br>(1.42, 2.30) | 2.73<br>(2.05, 3.64) |
|  | Mild | 1.50<br>(1.16, 1.94) | 1.86<br>(1.47, 2.37) | 2.32<br>(1.86, 2.90) | 2.86<br>(2.28, 3.58) |
|  | Moderate | 1.57<br>(1.09, 2.27) | 2.02<br>(1.49, 2.76) | 2.32<br>(1.77, 3.03) | 3.08<br>(2.45, 3.89) |
|  | Severe | 2.56<br>(1.59, 4.14) | 3.04<br>(2.04, 4.52) | 2.83<br>(2.01, 3.97) | 3.94<br>(3.09, 5.03) |

### Primary snoring and uncontrolled hypertension

To test for potential effects of primary snoring on uncontrolled hypertension, a similar analysis was applied to all participants with AHI <5 events/h. After excluding participants with an AHI ≥ 5 events/h, 4,529 (37%) participants remained, within which there were 607 (13·4%) cases of uncontrolled hypertension. The association between snoring duration and uncontrolled hypertension remained significant in this group. Participants who snored 5% (75^th^) of the night vs. those who did not 0·01% (5^th^ percentiles), had an 89% higher prevalence of uncontrolled hypertension (OR [95%CI], 1·89 [1·44, 2·46]), independent of age, sex, and BMI. The association between snoring and uncontrolled hypertension was non-linear, as shown in Appendix Figure 3.

## Discussion

This study showed that 15% of the population snore on average for more than 20% of the night and that ∼10% of participants without sleep apnea snore more than 12% of the night. The current findings also demonstrate that regular nightly snoring is associated with elevated blood pressure and uncontrolled hypertension, independent of OSA presence or severity. These findings provide important insight into the potential consequences of snoring on hypertension risk and highlight the need to consider snoring as part of clinical care and management of sleep problems, particularly in the context of hypertension management.

### Study strengths and limitations

There is considerable variability in night-to-night measures of OSA severity^17-19^. Thus, repeated measures over multiple months as performed in the current investigation provides substantially greater precision around estimates of OSA severity and snoring than previous studies that have relied on single night recordings. Moreover, the large-scale dataset surpasses all previous studies that have sought to investigate potential relationships between snoring and hypertension.

Objective data were also collected in the naturalistic home environment using simple low-cost consumer devices with established validation data. Multi-night measures in the home setting are more directly relevant to real-world risk exposure compared to data collected in sleep laboratory settings. Indeed, laboratory testing is typically derived from a single night in an unfamiliar environment which may confound both sleep and blood pressure measures.

There are several limitations of note. Firstly, there was a lack of assessment for clinical covariates that may confound the association between snoring duration and blood pressure. For example, smoking and alcohol consumption are associated with greater incidence and intensity of snoring and may have confounded the association with hypertension^24,25^. Furthermore, participants were predominantly male which limits generalization and raises the need for caution when interpreting sex difference comparisons. Participants were also self-selected via their decision to purchase and regularly use the under-the-mattress sleep sensor and blood pressure monitor devices from which these data were derived. Consumer choices could reflect concerns about sleep and blood pressure, potentially contributing to a bias towards overestimation of snoring prevalence. On the other hand, consumer engagement could potentially be biased towards those who are more engaged in their own health. Without any agreed standards as to how snoring should be assessed, meaningful comparisons of snoring prevalence across studies are problematic. For example, in our study, ∼15% of the population snored on average more than 20% of their estimated sleep time. In the Busselton study 22% of the studied population snored more than 50% of the total sleep time^15^. Standardized algorithms and methods for snoring assessment are clearly needed for meaningful comparisons of snoring prevalence across studies. Importantly, snoring algorithm design and cut-off choices should be informed by the associated adverse health impacts of snoring such as hypertension as reported in the current investigation.

It is also unclear how effectively the under-mattress sensors detect snoring from the participant and not a bed partner. This may have led to imprecise detection of snoring and influenced the estimated risk for uncontrolled hypertension associated with snoring. Finally, the under-the-mattress sleep sensor does not evaluate snoring intensity, which has previously been shown to be associated with increased next morning blood pressure^6^. Thus, evaluation of snoring loudness is also likely to be beneficial. Similarly, recent evidence suggests that snoring acoustic characteristics may be useful markers to detect the site(s) of upper airway collapse in people with OSA^26^. Analysis of snoring acoustic characteristics and intensity from under-the-mattress sensors, or potentially bed-side devices, therefore, warrants further investigation in OSA and in snorers.

### Clinical implications, and future research

The identification of associations between blood pressure and uncontrolled hypertension with snoring is consistent with previous much smaller studies and those reliant on subjective snoring assessments^6,7,27^. The moderating effect of age and BMI on the associations between snoring duration and hypertension is also consistent with prior OSA literature. Previous studies suggest that downstream effects of OSA *per se* on adverse health outcomes may be stronger in younger participants and potentially influenced by protective hypoxic pre-conditioning effects in older people with OSA^27,28^. Bixler and colleagues^28^ found that the association between OSA severity (measured using the AHI) and hypertension was stronger in younger and non-obese participants. Similarly, we found moderating effects of age and BMI, whereby snoring duration was more strongly associated with hypertension in younger and non-obese participants. In the same study, Bixler and colleagues^28^ did not find a moderation effect of BMI in the association between self-reported snoring and incident hypertension, which may be due to the subjective assessment of snoring. Nonetheless, the findings of this study indicate that simple non-invasive monitoring of snoring may be informative. Similarly, we have previously shown that AHI and inter-night variability of AHI estimated using the same under-the-mattress device are independently associated with hypertension risk^20^. Together, these findings indicate that non-invasive in-home monitoring over multi-nights may be clinically useful to better detect snoring and OSA, their potential adverse cardiovascular consequences, and to inform therapeutic decision making including in primary care.

The finding that the association between increased blood pressure and snoring, independent of OSA severity, has a similar effect size to the association between OSA severity and blood pressure in existing epidemiological trials supports that snoring may be an important mechanism that contributes to hypertension. For example, in the Sleep Heart Health Study, severe OSA (AHI > 30) vs no-OSA (AHI < 1·5 events/h) was associated with a 3·4 mmHg increase in diastolic blood pressure, independent of age, sex and BMI^29^. In this study, regular nightly snoring (4^th^ quartile vs. 1^st^ quartile) was associated with up to 4·5 mmHg higher diastolic blood pressure after adjustment for age, sex BMI and OSA severity. While uncontrolled factors may explain some of this effect (e.g., smoking or alcohol), snoring could also represent an independent mechanistic pathway via which chronically obstructed breathing during sleep contributes to elevated blood pressure. For example, via intrathoracic pressure effects on baroreceptor mediated control of systemic (and potentially pulmonary) blood pressure, or potentially via carotid baroreceptor effects from mechanical vibration emanating from the pharyngeal airway ^6,7,9-11^.

The elevated hypertension prevalence in people with primary snoring (AHI < 5 events/h) was comparable to other OSA severity categories. This finding further suggests that hypertension risks associated with snoring may be independent from OSA and potentially more strongly related to chronically obstructed breathing itself rather than blood gas or sleep disturbance effects. These results are concordant with previous studies where associations between snoring loudness and elevated morning blood pressure in patients with an AHI < 15 events/h were detected^6,7^. Some of this effect could potentially be explained, at least in part, by largely arbitrary definitions around hypopnea events, which do not capture obstruction severity or duration and may exhibit relatively weak relationships with snoring measures. Nonetheless, it may also reflect a specific risk associated with snoring itself^6,7,9-11^. Low level continuous positive airway pressure has been shown to significantly reduce snoring in participants without OSA^30^. Whether a reduction in snoring is associated with a reduction in blood pressure and other cardiovascular risk factors clearly warrants further investigation.

In summary, relatively long-term nightly snoring assessments indicate that snoring is highly prevalent in the adult community and is associated with an ∼20 to 80% increase in hypertension prevalence, independent of OSA severity. High hypertension prevalence was also observed for people with a high proportion of the night spent snoring, even without sleep apnea (AHI<5 events/h). These findings provide important insight into the potential consequences of snoring to hypertension. Thus, further investigation is warranted to determine whether therapeutic interventions directed towards snoring can reduce hypertension, one of the leading risk factors for cardiovascular disease and mortality.

## Data Availability

Deidentified data that support the findings of this study, including individual data, are available from the corresponding author upon request subject to ethical and data custodian (Withings) approval.

## Acknowledgments

DJE is supported by a National Health and Medical Research Council of Australia (NHMRC) Senior Research Fellowship (1116942) and an Investigator Grant (1196261).

## Author contribution

DJE, BL, GN, SA and PC developed the study concepts and aims. BL, and GN performed the data extraction. BL, DJE and GN performed the data analysis. DJE, BL, GN, HS and JM drafted the manuscript. All authors provided important insight on data analysis, interpretation and contributed to the final version of the manuscript. All authors had full access to all the data in the study and accept responsibility to submit for publication

