## Supplementary for "Regular snoring is associated with uncontrolled hypertension: A longitudinal objective assessment of nightly snoring and blood pressure"

Mark Oliphant Building, Level 2, Building A, 5 Laffer Drive, Bedford Park 5042

[1] Adelaide Institute for Sleep Health and FHMRI Sleep Health, College of Medicine and Public Health, Flinders University, Adelaide, Australia

[2] College of Science and Engineering, Flinders University, Adelaide, Australia

[3] Centre Interdisciplinaire du Sommeil, Paris, France

**Snoring and blood pressure**

There was an interaction between snoring duration and age ($p = 0.008$) in the association between snoring and diastolic blood pressure, where the effect of snoring duration on diastolic blood pressure was more pronounced in younger participants (Figure 1). Furthermore, the association between snoring duration and diastolic blood pressure was higher (e.g., steeper increase; p-value for interactions, $p < 0.001$) for participants with a BMI ≥ 30 compared to participants with a BMI < 30 kg/m².

Similarly, there was a significant interaction with BMI ($p < 0.001$), but not age ($p = 0.600$) in the association between snoring duration and systolic blood pressure. There was also an interaction between snoring duration and sex (Supplementary Table S1, $p < 0.001$) in the association with systolic blood pressure. Conversely to diastolic blood pressure, participants with a BMI < 30 kg/m² (Figure 1) and women (Supplementary Table S1) had a higher increase in systolic blood pressure associated with snoring duration than obese participants and men.

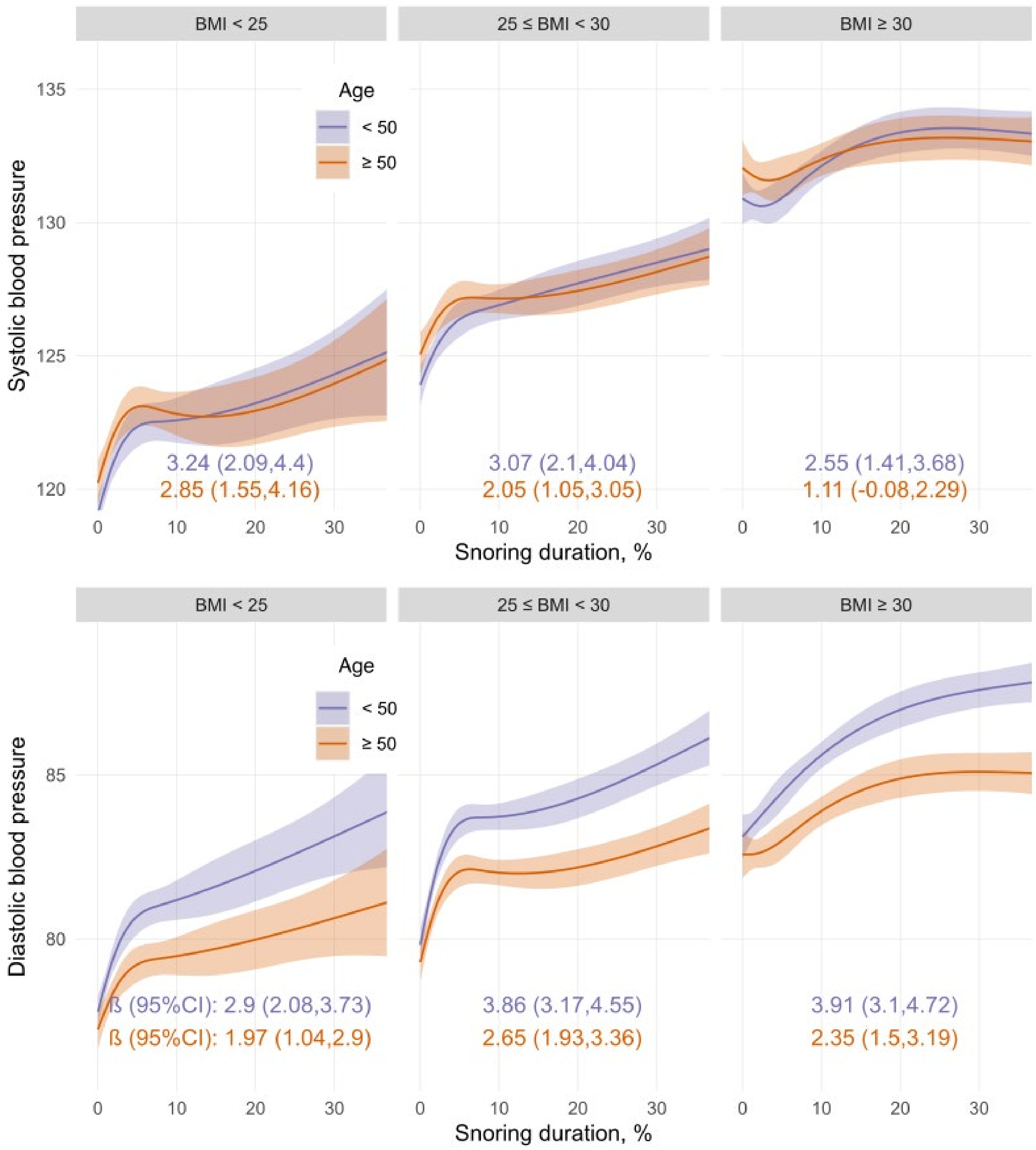


**Figure S1.** *Associations between snoring duration with systolic and diastolic blood pressure, modelled using 4 knots restricted cubic spline for snoring duration and an interaction with age categories in years (median split) and BMI categories (kg/m$^2$). β (95%CI) represents the difference between the 5% and the 75% percent of the snoring duration distribution. Note that the 5% and 75% percent were determined separately for each BMI category.*

***Table S1****: Association between snoring and systolic blood pressure modelled with an interaction term between snoring and BMI categories, and an interaction term between snoring and sex. Results represents the blood pressure difference β (95%CI) between the 75$^{th}$ vs 5$^{th}$ percentile of the snoring distribution. Models are adjusted for age and apnea-hypopnea-index.*

| | Men | Women |
|---|---|---|
| BMI < 25 | 2.71 (1.61, 3.81) | 3.90 (2.06, 5.74) |
| 25 < BMI < 30 | 2.17 (1.34, 3.00) | 5.20 (3.53, 6.87) |
| BMI > 30 | 1.47 (0.44, 2.50) | 5.33 (3.50, 7.15) |

**Snoring and hypertension**

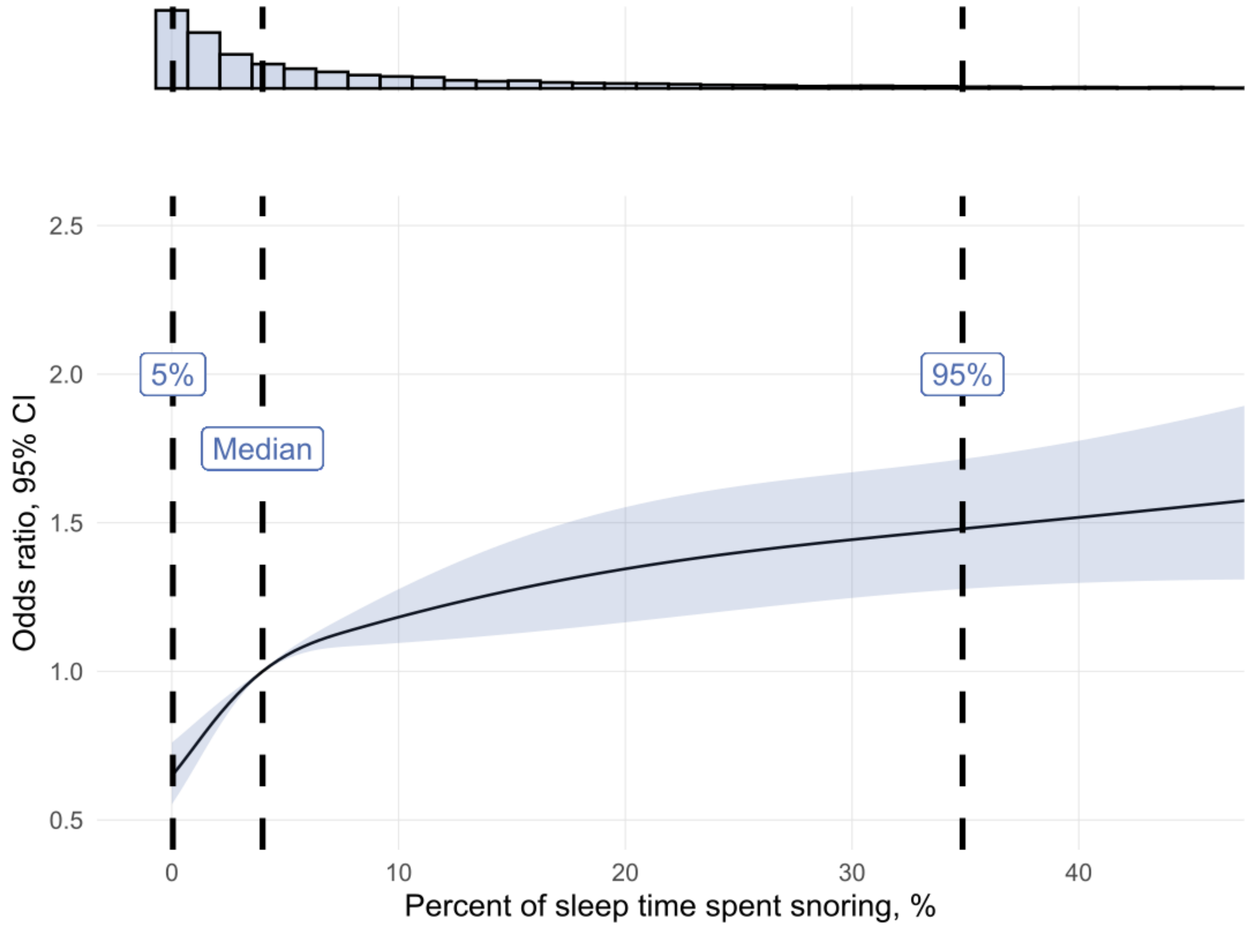


***Figure S2.*** *Associations between snoring (as a % of total sleep time) and hypertension risk.*

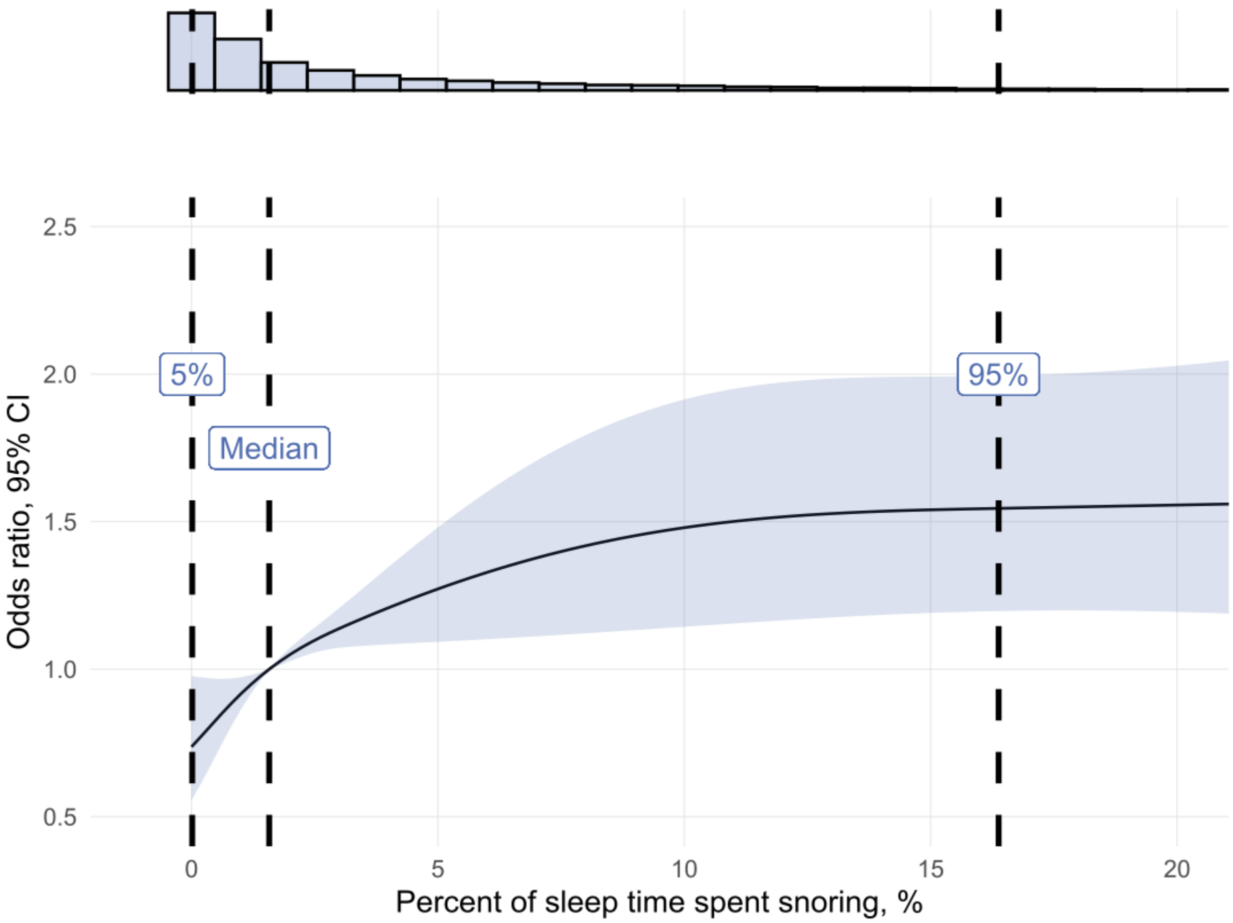


***Figure S2.*** *Associations between snoring (as a % of total sleep time) and hypertension risk in participant with no sleep apnoea (apnoea-hypopnea index < 5).*
